# Investigating duration and intensity of Covid-19 social-distancing strategies

**DOI:** 10.1101/2020.04.24.20078022

**Authors:** C. Neuwirth, C. Gruber, T Murphy

## Abstract

The exponential character of the recent Covid-19 outbreak requires a change in strategy from containment to mitigation. Meanwhile, most countries apply social distancing with the objective to keep the number of critical cases below the capabilities of the health care system. Due to the novelty and rapid spread of the virus, an a priori assessment of this strategy was not possible. In this study, we present a model-based systems analysis to assess the effectiveness of social distancing measures in terms of intensity and duration of application. Results show a super-linear scaling between intensity (percent contact reduction) and required duration of application to have an added value (a lower number of fatalities). This holds true for an effective reproduction of *R* > 1 and is reverted for *R* < 1. If R is not reduced below 1, secondary effects of required long-term isolation are likely to unravel the added value of disease mitigation. If an extinction is not feasible, we recommend moderate social-distancing that is well balanced against capability limits of national health-care systems.

## Introduction

This article is written in mid-April 2020 where globally the number of confirmed COVID-19 cases is above 2.5 million resulting in over 170,000 deaths [1]. Due to these large numbers, the initial approach of containment, i.e. tracing contacts of patients with laboratory-confirmed infection [2], is not applicable anymore and may even have unintended consequences of hampering effective healthcare delivery [3]. Instead, the majority of countries decided to use community interventions, like cancellation of events, general social distancing and travel restrictions [4].

These community mitigation measures reduce transmission, hence flatten the curve and push the peak of new infections further into the future which eventually helps preventing an epidemic peak that overwhelms health-care systems [5]. The need for mitigation measures is also evident from the actions taken by European countries who implemented interventions including the closure of schools and universities, banning of mass gatherings, and most recently, widescale social distancing including local and national lockdowns [6].

Numerous studies support this strategy [3] [4] [7] [8]. Applied in the long-term, however, school closure and home confinement will negatively affect children’s health [9] and the global economy, to name only two big drawbacks of these measures. In this light, it is of particular interest, for what duration these exceptional interventions must remain in place. According to recent estimates, we are probably at least 1 year to 18 month away from large-scale vaccine production [5]. Independent of the time it takes to develop a vaccine, the epidemic spread will also come to an end, if sufficient people have been infected to establish herd immunity. Studies on the effectiveness of the concept of disease mitigation with the objective to establish herd immunity shows some potential in the case of pandemic influenza [10].

In this study, we present an exploratory and model-based systems analysis that is aimed at investigating the application of social distancing strategies to Covid-19. Specific objectives of this research are: 1) to investigate the effectiveness of contact reduction policies with respect to intensity and duration and 2) to estimate the amount of time to establish herd immunity by considering the national health care systems of Austria and Sweden, which are very different in terms of critical care capabilities (Austria: 21.8 beds Sweden: 5.8 beds per 100k population, respectively) [11]. A detailed description of model equations, assumptions as well as uncertainty of currently available data are presented in the following section. Data uncertainty is addressed by the analysis of alternative scenario runs to enhance robustness of model results. In a concluding section, we compare our results to similar studies, discuss current limitations of data availability and give recommendations based on exploratory results.

## Method

### Adapted SIR model

The current scenario of novel pathogen emergence includes considerable uncertainty [12]. This means that a reliable scientific evidence base on Covid-19 is yet to be established. Under these preconditions, the use of models for exploratory rather than predictive purposes is more appropriate [13]. Accordingly, the simulation model presented in this study was designed to identify and systematically explore important qualitative behavior of this dynamic system that remains unchanged irrespective of parameter variations. An adaptation of the popular-infected-recovered (SIR) model turned out to be most suitable for this purpose (see Fig. 1). In order to meet the specific requirements of a simulation model on Covid-19 mitigation, the structure of the original model was adapted accordingly. For instance, pathological findings of Covid-19 indicate that there is a considerable number of cases that develop mild or no symptoms [14]. To account for this characteristic, we separated the infected population into those that are asymptomatic and those that are not, which in the latter case leads to isolation or hospitalization. The asymptomatic infected get resistant without prior isolation.

**Fig 1.**
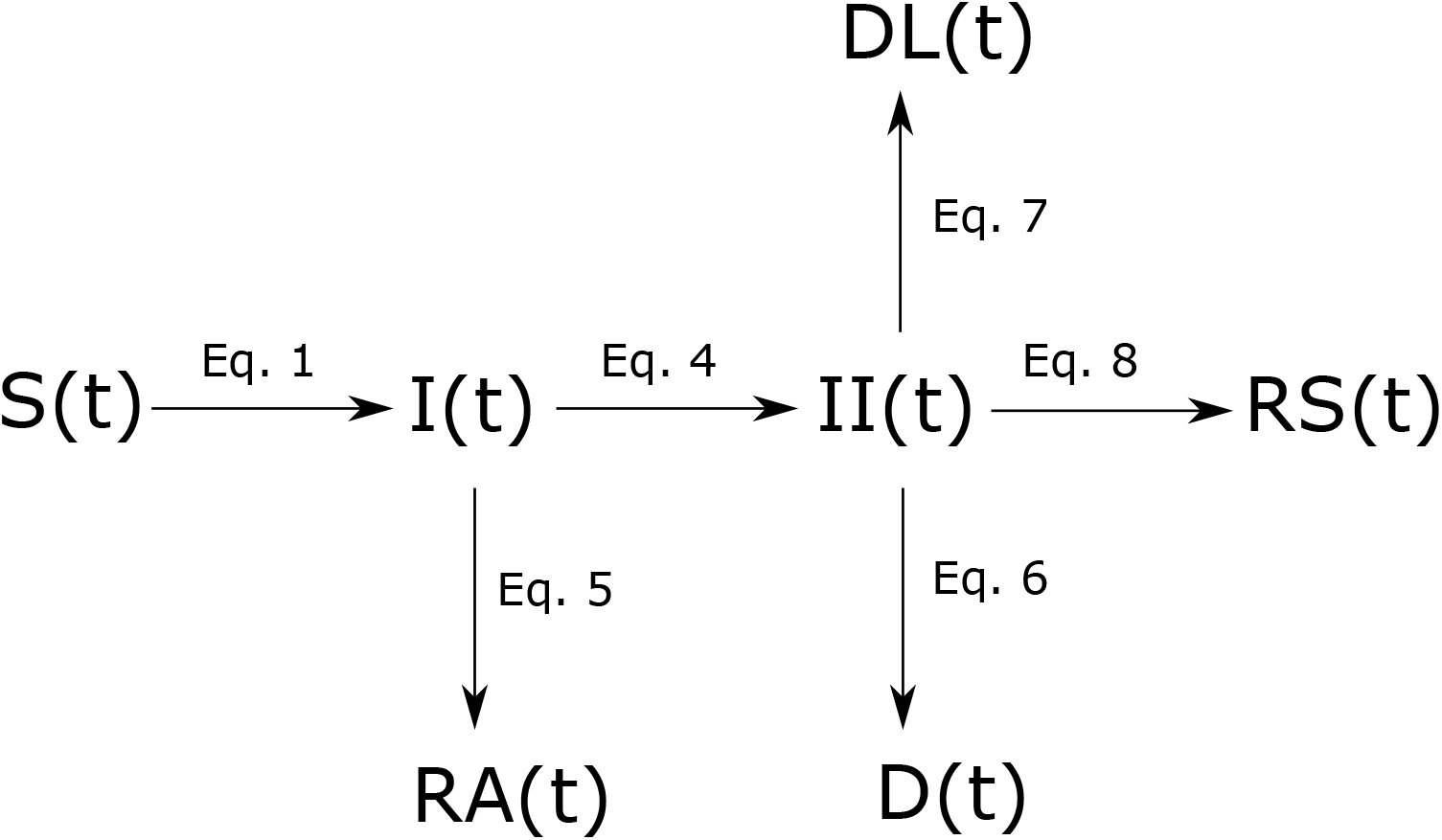
**Schematic diagram of the adapted SIR model:** susceptible S(t), infected - infection unknown I(t), infected in isolation II(t), resistant symptomatic RS(t), resistant asymptomatic RA(t), deaths D(t), deaths caused by lack of ICU DL(t), compare equations 1 - 8

Exponential growth in numbers of infected poses a challenge to health care facilities. In Italy, specialists are already considering denying life-saving care to the sickest and giving priority to those patients most likely to survive [15]. This will inevitably cause potentially avoidable deaths. In the model, deaths caused by a lack of intensive care is considered independently.

The calculation of population quantities in respective compartments (see Fig. 1) is in line with the logic of the standard SIR model. The model makes the simplified assumption that initially everyone in the total population *N* is susceptible. The number of susceptible is reduced over time by infections as

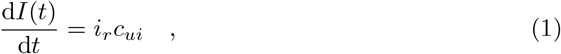

where *i*_*r*_ is the infection rate (rate of contacts between uninfected and infected that result in infections) and *c*_*ui*_ is the number of contacts between infected and uninfected, which is calculated as

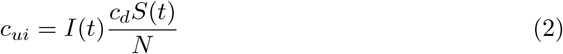

where c_*d*_ is the personal contacts per day, *S*(*t*) is the susceptible at time t and I(t) is the number of unknown infections. To take account for a lower infection rate of asymptomatic infected, the unknown infections I(t) in equation 2 is substituted by

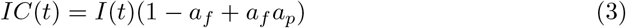

with IC(t) being the unknown infected corrected for asymptomatic infected, *a*_*f*_ the fraction of asymptomatic among infected and *a*_*p*_ the asymptomatic population’s potential to infect.

The flows from compartment I(t) - i.e. asymptomatic cases getting resistant 5 and isolation of symptomatic infected 4 –are calculated by

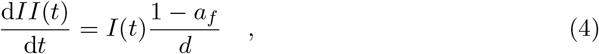

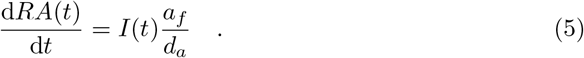

Parameter d is the time between infection and isolation (corresponds to the duration of infectiousness) and *d*_*a*_ is the duration of asymptomatic infection. The flows from compartment II(t) are given by

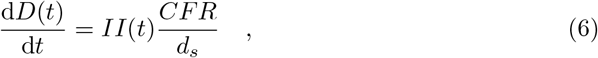

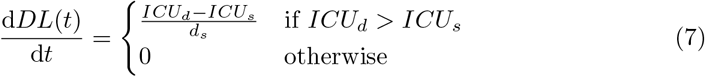

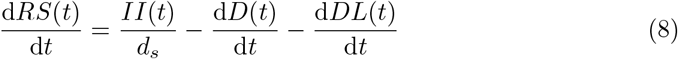

where parameter *d*_*s*_ is the duration of distinct symptomatic sickness, CFR is the case fatality rate and *ICU*_*d*_ and *ICU*_*s*_ is the intensive care demand and supply respectively.

The intensive care demand *ICU*_*d*_ is calculated by taking the critical fraction of infected in isolation II(t). The fraction of infected who are admitted to intensive care is denoted as *c*_*f*_ (see Table 1).

**Table 1.** Model parameters

| Parameters | uncertainty | Value(s) |
| --- | --- | --- |
| basic reproductive numbers ($R_0$) | high | 1.4 [16]<br>2.1 [30]<br>3.2 [31] |
| percentage of infected population that are asymptomatic ($a_f$) | high | 17.9% [19]<br>50% [21] |
| asymptomatic population's potential to infect ($a_p$) | high | 50% [32]<br>100% |
| number of available ICU beds/100000 inhabitants | medium | AUT 21.8<br>SWE 5.8<br>[11] |
| duration of distinct symptomatic sickness ($d_s$) | medium | 7 days [14] |
| duration of asymptomatic infection ($d_a$) | medium | 14 days (assumed to be similar to symptomatic) |
| initial infected population of total population ($I_0$) | model input | $S_0 \times 10^{-6}$ |
| ratio of confirmed cases that need intensive care ($c_f$) | high | 5% [23] 2.2% [26] |
| case fatality rate ($CFR$) | high | 0.7% [28] |
| social contact reduction in percent ($c_r$) | model input | |
| duration of precautionary measures ($d_m$) | model input | |

### Model inputs and exploration

Despite the novelty of Covid-19, the body of literature on key parameters like basic reproduction, case fatality rates and proportion of asymptomatic cases is quite substantial and growing. The wide range of suggested parameter values, however, poses a considerable challenge to model parametrization. For instance, estimates of the basic reproduction number R0 vary within a range from 1.4 [16] to 4.71 [17]. Part of this variation is explained by geographic variation of population densities as well as by heterogeneous social and cultural habits [18]. Moreover, there is uncertainty in the percentage of asymptomatic cases. The outbreak in a smaller isolated population is an opportunity to derive representative numbers by applying comprehensive and repeated laboratory testing. One such example is the outbreak of Covid-19 on board of the Diamond Princess cruise ship. However, given that most of the passengers were 60 years and older, the nature of the age distribution may lead to underestimation of asymptomatic cases if older individuals tend to experience more symptoms [19]. The assumed age dependence of asymptomatic infection is supported by a screening of pregnant woman admitted for delivery in New York-Presbyterian Allen Hospital between March 22 and April 4, 2020 (n=215) that show an asymptomatic fraction at presentation of 87.9% among 33 patients who were tested positive for SARS-CoV-2 [20] In a normal population ratios of about 50% asymptomatic carriers of Covid-19 are expected ([21]). The question whether or not asymptomatic carriers are able to infect others is still controversial (e.g. [22]).

The severity of the disease does also play an important role in estimating the ratio of critically ill patients who need intensive care. According to Chinese statistics, 5% of positively tested patients are admitted to intensive care [23]. This number was adopted by the World Health Organization [24] and other studies (e.g. [25]), whereas national statistics show significant deviations; e.g. 9 to 11% in Italy [15] and 2.2% in Austria [26]. A potential explanation for these considerable differences is that in Italy a lot of the older population were infected [27].

The specific age distributions of affected communities may also show some biasing effect on estimated case fatality rates. Another factor that contributes to regional differences in case fatality is the occupation or over-occupation of available intensive care beds (ICU beds). In a few instances, national critical care capabilities are exceeded by the number of critically ill patients (e.g. Italy and France), which drastically elevates fatality rates. By contrast, the true case fatality rates are lower if theoretically all cases were found by testing the entire population. Accordingly, a lower case fatality rate (CFR) was reported by countries who were effective in extensive testing and maintaining the prevalence of critical cases below critical care capabilities like South Korea [28]. A higher CFR was reported by countries who refrain from extensive testing and/or are overwhelmed by the pace of new infections like Iran, Italy and others [29]. In the model, we use the more reliable South Korean figures and simulate the additional fatalities due to the critical care limit based on capability limits of national critical care units (see Eq. 6 and Eq. 7).

Among the parameters in Table 1, the basic reproduction number R0 is the only parameter without explicit representation in the model equations. This parameter is the number of secondary cases, which an infected person produces in a completely susceptible population [33]. In the absence of asymptomatic infections, R0 in the model is equivalent to the arithmetic product of d the time between infection and isolation, *c*_*d*_ the personal contacts per day and i_*r*_ the infection rate.

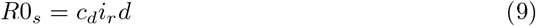

To account for asymptomatic infection, *R*0_*s*_ is modified by the specific duration of infectiousness (*d*_*a*_) and infection potential (*a*_*p*_) of asymptomatic infected. This can be expressed as

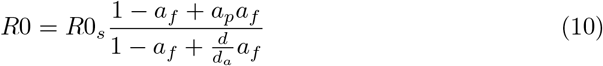

The numerator scales *R*0 by the asymptomatic populations’s infectiousness, whereas the denominator scales *R*0 by the asymptomatic population’s duration of infectiousness.

In response to an epidemic disease outbreak, measures are implemented to reduce contacts between people thereby reducing the reproduction of the disease. This is implemented in the model by lowering the personal contacts per day *c*_*d*_. We refer to this modified reproduction as effective reproduction number R. The choice of appropriate scenarios is based on parameter uncertainty and model sensitivity.

Sensitivity analysis indicate a linear response in model output to variations in CFR and *c*_*f*_, and interestingly non-linear effects in response to variations in R, *a*_*f*_ and *a*_*p*_. Accordingly, the latter variables were selected as scenario parameters (see Table 2).

**Table 2.** Scenario runs.

| Nr | Scenario name | Parameters | Type |
| --- | --- | --- | --- |
| 1 | symptomatic, high basic reproduction | $a_f=17.9\%$,<br>$a_p=50\%$,<br>$R_0=3.2$ | prolonged |
| 2 | symptomatic, medium basic reproduction | $a_f=17.9\%$,<br>$a_p=50\%$,<br>$R_0=2.1$ | prolonged |
| 3 | symptomatic, low basic reproduction | $a_f=17.9\%$,<br>$a_p=50\%$,<br>$R_0=1.4$ | prolonged |
| 4 | asymptomatic, high basic reproduction | $a_f=50\%$,<br>$a_p=100\%$,<br>$R_0=4.26$ | prolonged |
| 5 | asymptomatic, medium basic reproduction | $a_f=50\%$,<br>$a_p=100\%$,<br>$R_0=2.8$ | prolonged |
| 6 | asymptomatic, low basic reproduction | $a_f=50\%$,<br>$a_p=100\%$,<br>$R_0=1.86$ | prolonged |
| 7 | medium basic reproduction, high need for intensive care, Austria | $a_f=17.9\%$,<br>$a_p=50\%$,<br>$R_0=2.1$,<br>$ICU_s=21.8$,<br>$c_f=5\%$ | intermittent |
| 8 | medium basic reproduction, high need for intensive care, Sweden | $a_f=17.9\%$,<br>$a_p=50\%$,<br>$R_0=2.1$,<br>$ICU_s=5.8$,<br>$c_f=5\%$ | intermittent |
| 9 | high basic reproduction, high need for intensive care, Austria | $a_f=50\%$,<br>$a_p=100\%$,<br>$R_0=2.8$,<br>$ICU_s=21.8$,<br>$c_f=5\%$ | intermittent |
| 10 | high basic reproduction, high need for intensive care, Sweden | $a_f=50\%$,<br>$a_p=100\%$,<br>$R_0=2.8$,<br>$ICU_s=5.8$,<br>$c_f=5\%$ | intermittent |
| 11 | low basic reproduction, low need for intensive care (Felix Austria) | $a_f=17.9\%$,<br>$a_p=50\%$,<br>$R_0=1.4$,<br>$ICU_s=21.8$,<br>$c_f=2.2\%$ | intermittent |

Moreover, prolonged and intermittent social distancing [16] was applied in respective scenarios (see Table 2). Whereas prolonged social distancing is defined by constants *c*_*r*_and *d*_*m*_, intermittent social distancing is implemented by dynamic adaptation of contact reduction *c*_*r*_ during simulation runtime dependent on the amount of ICU beds available. If more than 70% of ICU beds are vacant, measures are loosened (daily change of *c*_*r*_ −20% until initial *c*_*d*_ is reached), whereas measures are tightened (daily change of *c*_*r*_ +20% until *c*_*d*_ = 0 is reached) in case less than 30% of ICU beds are available. The linear adjustment in measures is based on the assumption that a complete lockdown (*c*_*r*_ = 100%, *c*_*d*_ = 0) can be removed or implemented within 5 days.

## Results and discussion

### Effectiveness of contact reduction

An increase in asymptomatic cases will overall increase the potential of the infected population to infect susceptible people, i.e. increase the basic reproduction R0, provided asymptomatic and symptomatic cases have a similar potential to infect. This is due to the extended duration asymptomatic cases remain undetected and thus infectious. The reduction of potentially infective contacts has the opposite effect and thus diminishes R0.

Moreover, the simulation shows that a lowering of the effective reproduction number flattens the curve and delays the peak of new infections, whereas an increase has the opposite effect (see Fig. 2). Consequently, social distancing flattens the curve of daily infections, while higher proportions of asymptomatic cases elevate the peak (provided *a*_*p*_ = 1 and *d* < *d*_*a*_).

**Fig 2.**
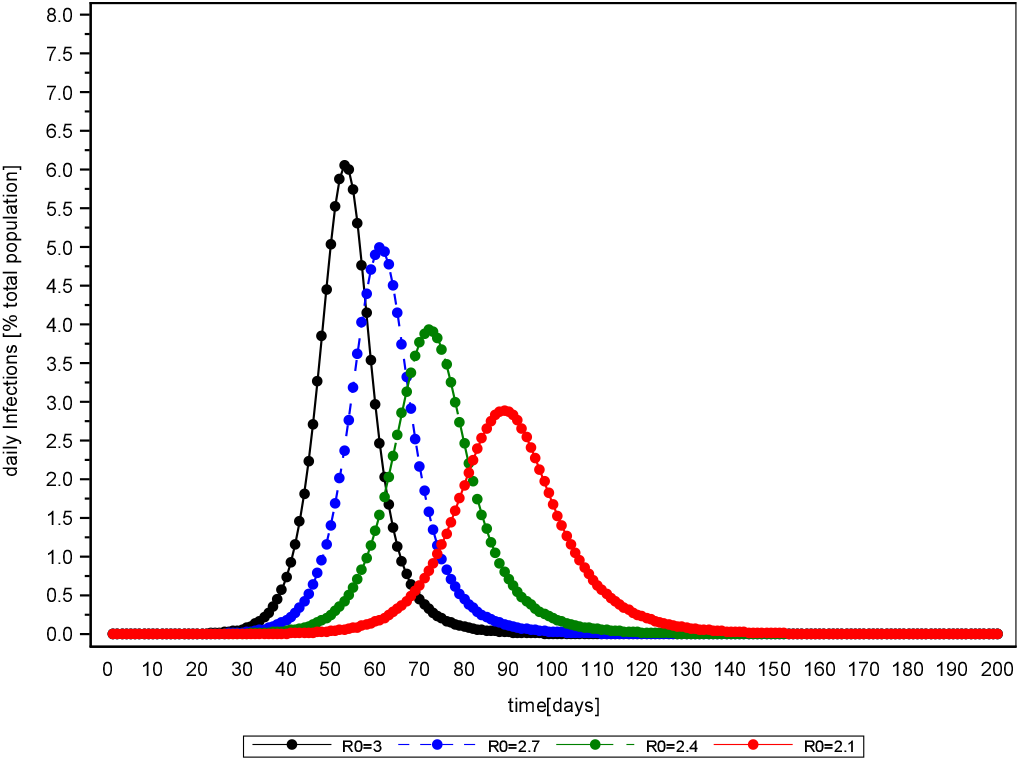
Delay effect of mitigation interventions (*a*_*f*_ = 0)

This flattening effect can be expressed analytically. The daily infections resemble a normal distribution, which is defined by a mean *µ* (days between outbreak and peak of daily infections) and a standard deviation *σ*. A lower R will lead to a higher *µ* (see Fig. 3) and a higher *σ* (see Fig. 4). Additionally, the number of initial infected people reduces *µ* (see Fig. 5), whereas *σ* is independent of it. These inverse relationships can be explained by mechanisms of viral spread. The effective reproduction of the disease at a given time t diminishes with every new infection that depletes the susceptible population (denoted as R(t)). Once R(t) drops below 1, the curve of new infections has passed its peak. In the case where initial effective reproduction R is low, the pool of susceptible individuals is slowly depleted (slow viral spread) and the peak at R(t)=1 is reached at a later point in time, which produces a flatter curver (high *µ* and *σ*).

**Fig 3.**
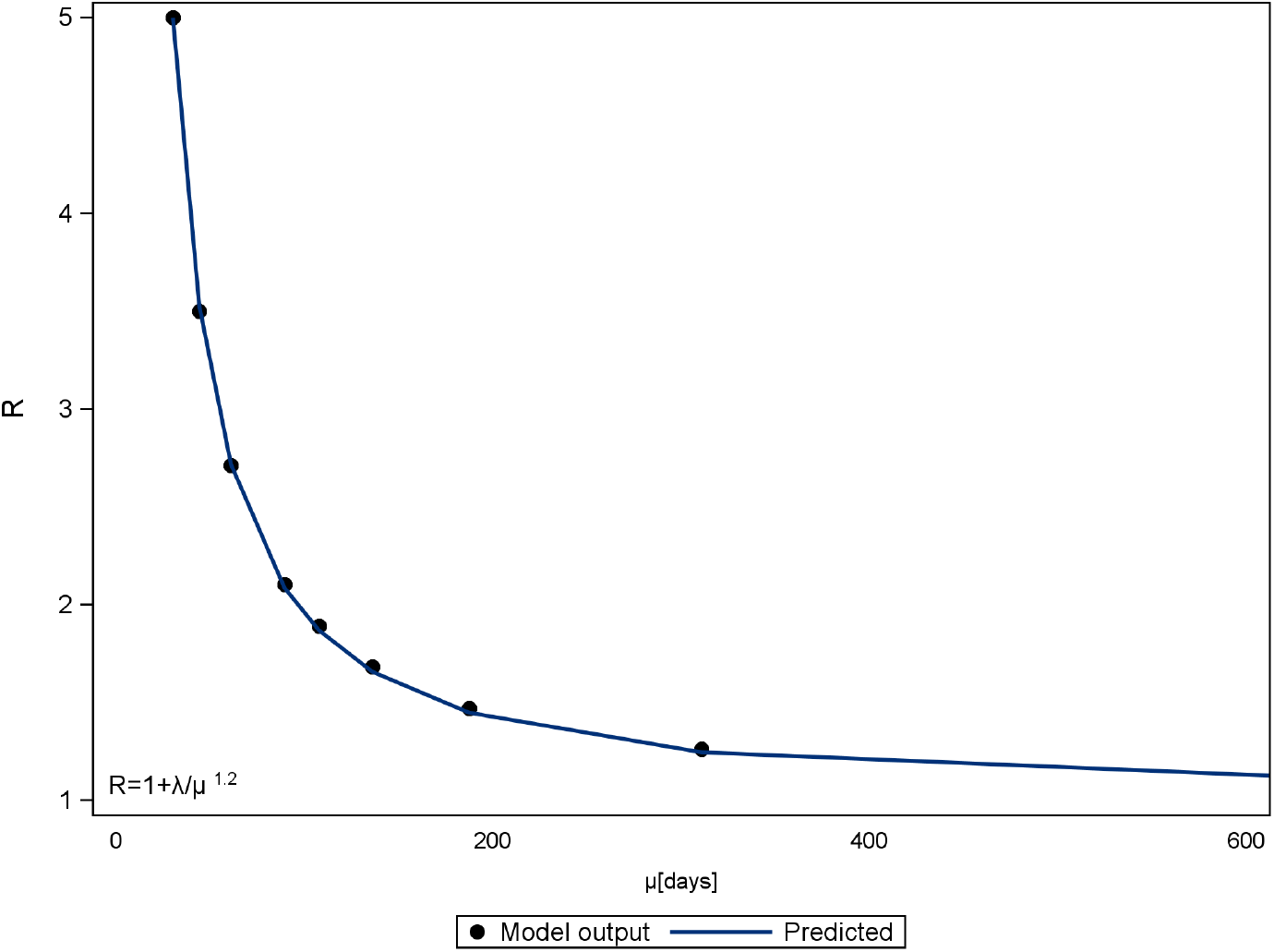
Relationship between effective reproduction R and peak occurrence *µ* in days after disease outbreak (*a*_*f*_ = 0). The parameter *λ* is calculated to 238

**Fig 4.**
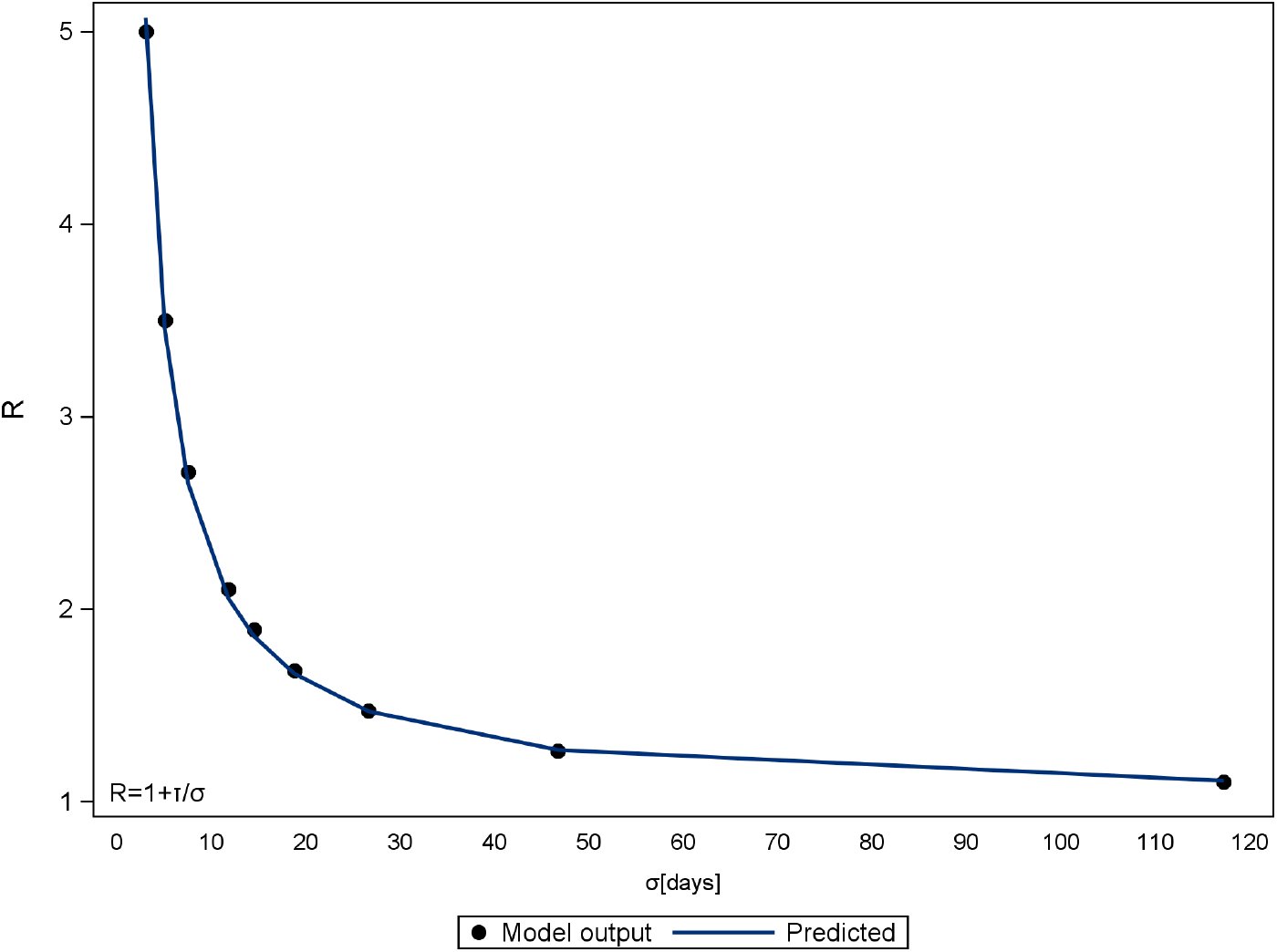
Relationship between effective reproduction R and standard deviation *σ*. (*a*_*f*_ = 0) The parameter *τ* is calculated to 12.46

**Fig 5.**
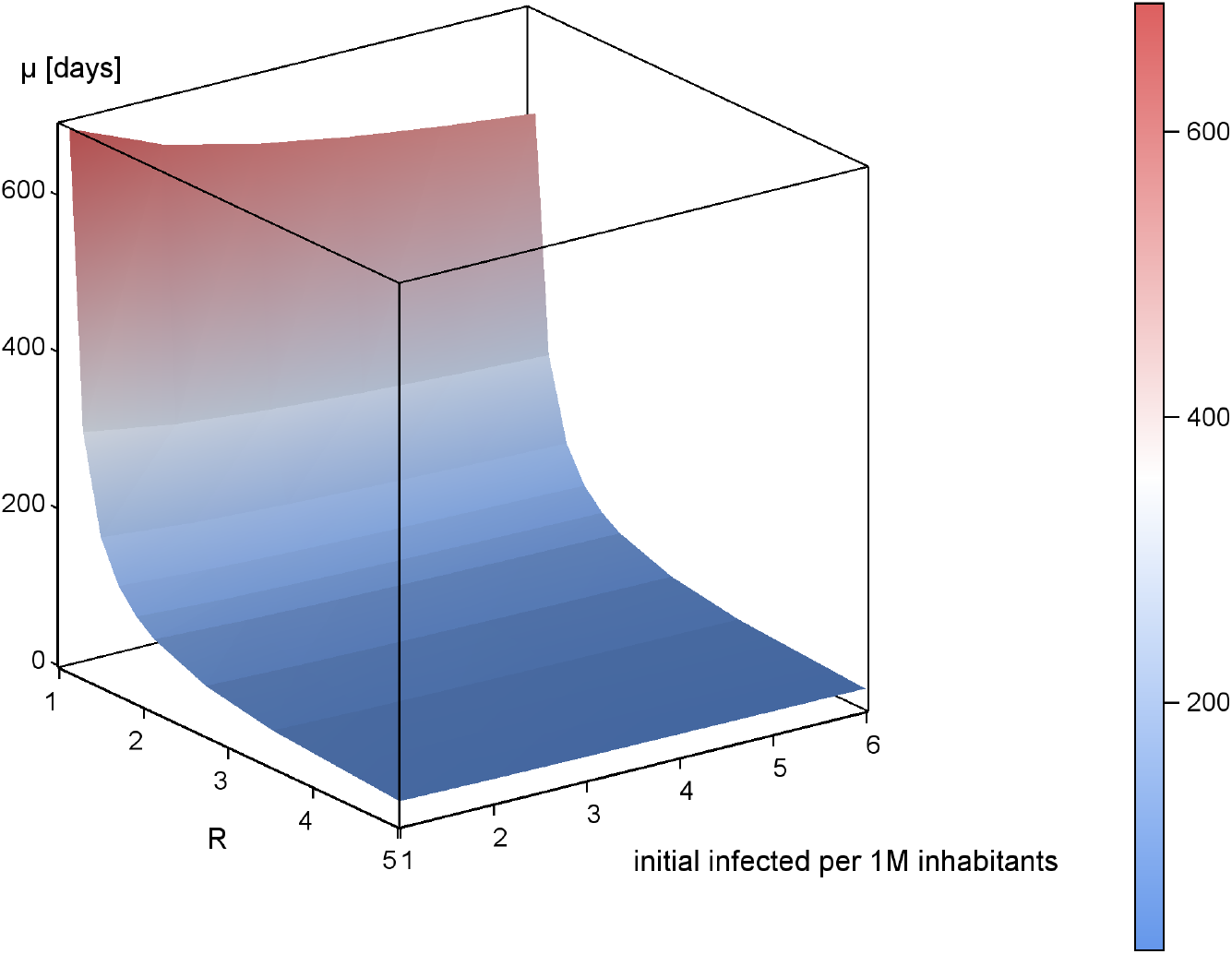
Relationship between effective reproduction, peak occurrence (*µ*) and number of infected at model initialization (*a*_*f*_ = 0).

These non-linear relationships have an important impact on the effectiveness of interventions. Social contact reduction and associated reduction in R push new infections further into the future. Hence, the more intense the social distancing measures in terms of contact reduction, the longer the duration needs to be to have an added value; i.e. a lower number of fatalities. In other words, the harder you break, the longer it takes.

For instance, 40% contact reduction needs to be applied for additional 600 days to outperform a 30% contact reduction in scenario 2 (see Fig. 6). The lower the basic reproduction in the scenarios, the larger the time lag associated with an intensification of social distancing (see Fig. 6, scenarios 1, 2, 4, 5 and 6). This is in line with above-mentioned relationships that show increased effects of R on *µ* and *σ* with lower R.

**Fig 6.**
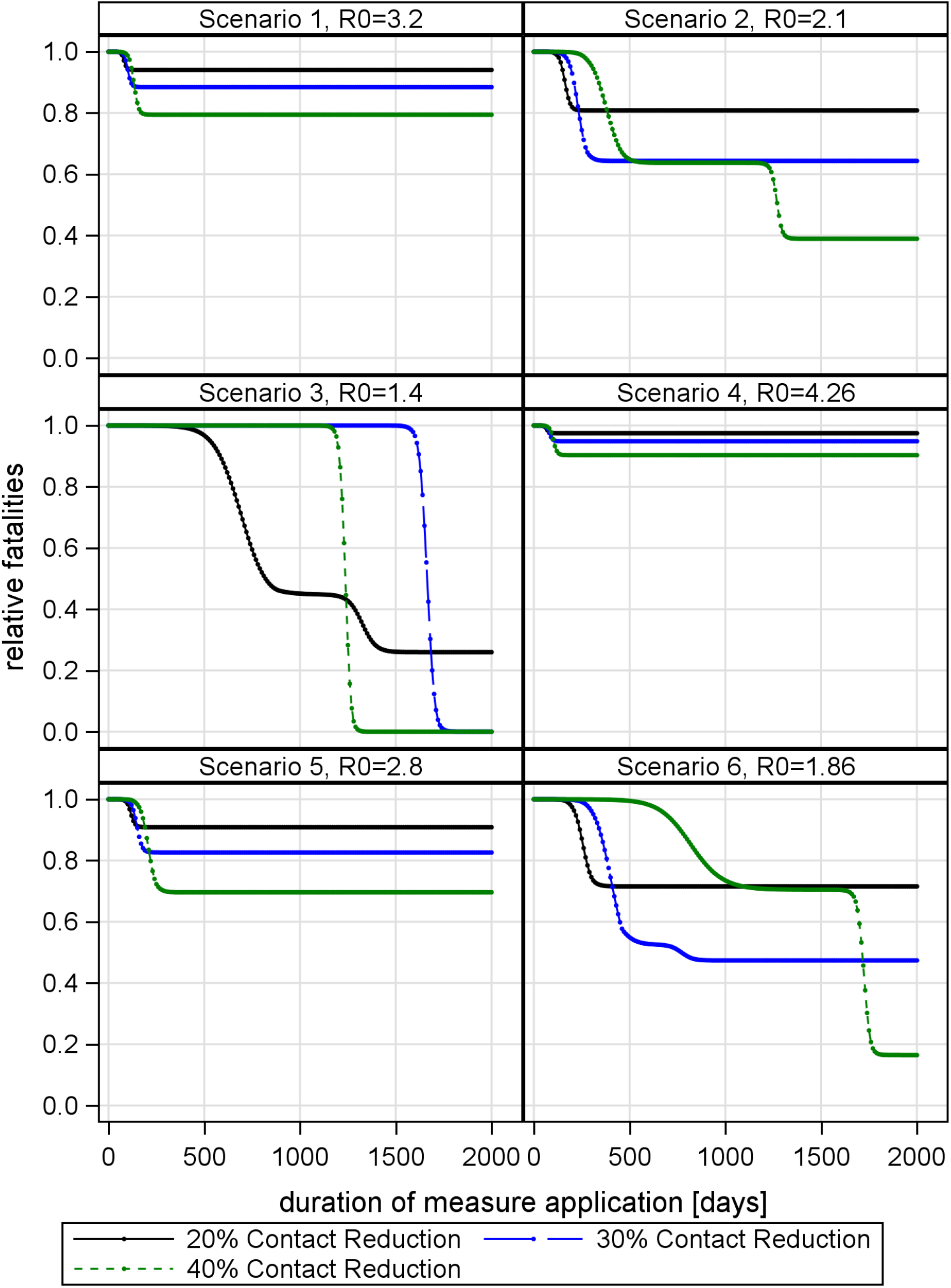
Relative fatalities (fatalities with contact reduction divided by fatalities without contact reduction, i.e. a relative fatality of 1 indicates that measures have no added value) plotted against social contact reduction in percent and duration of measure application.

Given the trade-offs associated with required long-term lockdown, the effectiveness of additional social distancing decreases with R close to 1. The secondary effects of lock down have not been modelled, but it is speculated that reductions in social contacts will increase mortality (e.g. social isolation and homicide; obesity and cardiovascular diseases etc.) making moderate contact reduction more adequate.

Interestingly, if social distancing is intense enough to drop R below one, a further increase in intensity reduces the needed duration of social distancing. For example, in scenario 3 (see Fig. 6), a 30% contact reduction as well as 40% contact reduction push R below one. The 40% contact reduction shows a positive effect (a lower number of fatalities) earlier than the 30% contact reduction. This is contrary to the case of *R* > 1 where the effectiveness of more intense measures is in danger to be unraveled by the super-linear increase in duration.

The curve flattening effect of social contact reduction also explains why drastic contact reduction may cause more deaths than mild contact reduction, if measures are applied for too short time. In the worst case, intense social distancing will hardly have any effect (see Fig. 7).

**Fig 7.**
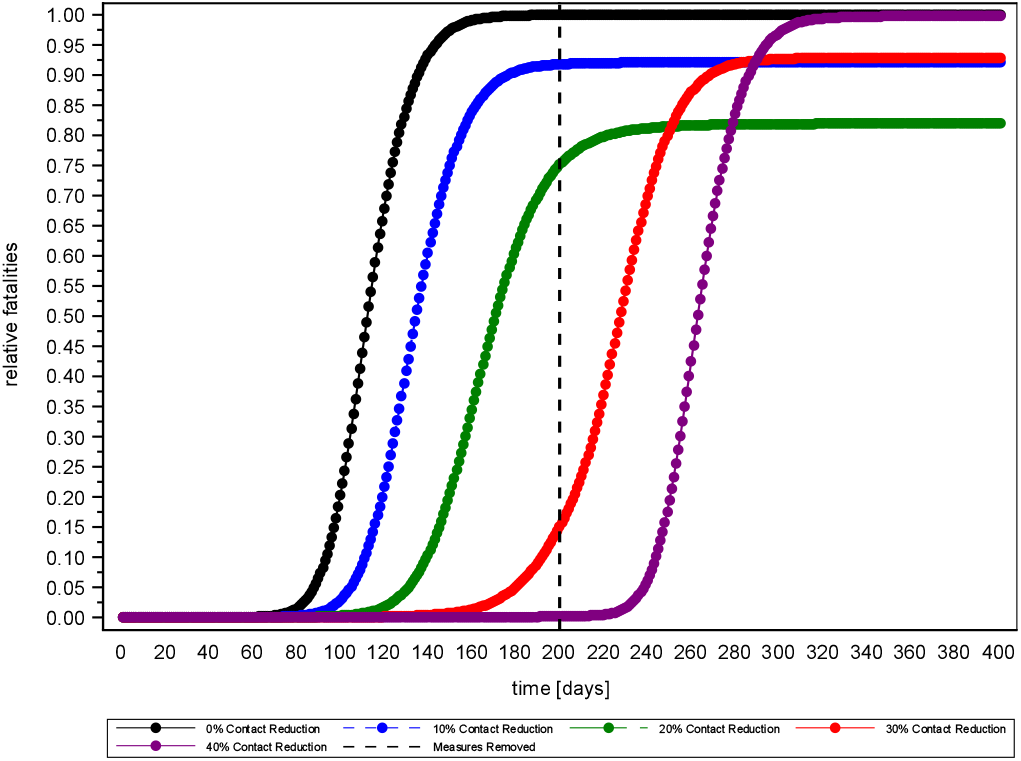
Relative fatalities (fatalities with contact reduction divided by fatalities without contact reduction, i.e. a relative fatality of 1 indicates that measures have no added value) in Scenario 2, (R0=2.1) with constant duration (200 days) and varying intensity of contact reduction.

### Duration to establish herd immunity

Intensity and duration are also closely related in the intermittent social distancing and herd immunity scenario (see Fig. 8). The strategic objective in this scenario is to keep the demand for ICU beds within the bounds of ICU supply until herd immunity is established.

**Fig 8.**
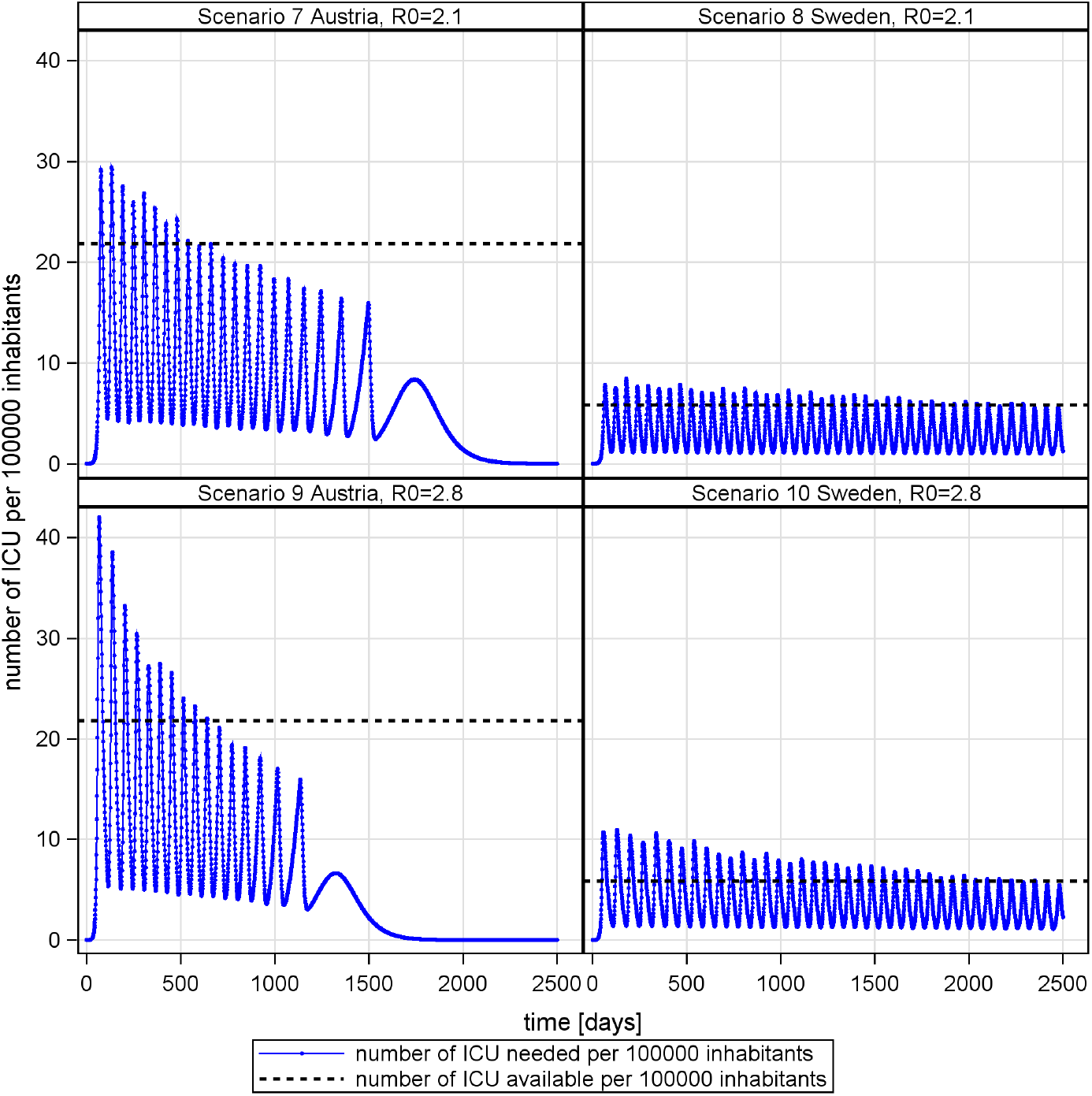
Policy based mitigation of new infections to meet capabilities of national health care systems.

In the simulation, the demand for ICU beds behaves like a damped oscillation (see Fig. 8). This is explained by the delay in the system, diminishing number of susceptible people and the negative feedback between number of available ICU beds and social contacts.

In the early phase of the outbreak, the number of patients exceeds the number of available ICU beds due to high reproduction potentials. Higher basic reproduction R0 results in additional over-occupation of ICU capabilities (see Fig. 8, scenario 9 and 10).

Moreover, the variation of available intensive care brings about a shift in the time needed to achieve the strategic objective of herd immunity (compare Sweden and Austria in Fig. 8). This relationship exhibits an almost linear scaling. Furthermore, results show that even under favorable conditions, social distancing and herd immunity strategies require extraordinary endurance.

In Austria, for instance, it is estimated that only 2.2% of confirmed cases are admitted to ICU [26]. Combined with Austria’s high performance health care system and low effective reproduction, the time to establish herd immunity is still estimated to be about 2 years (see Fig. 9). Given that the ICU beds are also needed for patients other than Covid-19, an even longer period has to be expected.

**Fig 9.**
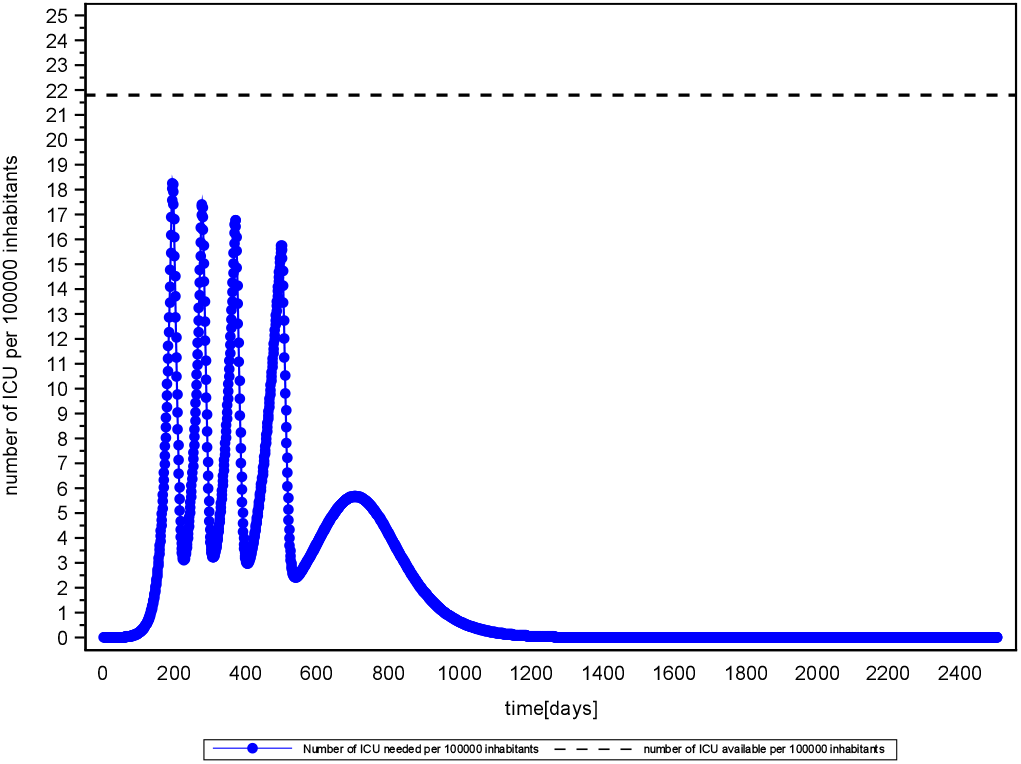
Mitigation and herd immunity strategy in a high performance health care system (Scenario 11 Austria, R0=1.4) and low rates of ICU admissions (2.2% of confirmed infected)

## Conclusion

In this article, we used methods of exploratory systems simulation to assess the effectiveness of social distancing measures in the mitigation of Covid-19. The simulated behavior is governed by a non-linear relationship between the intensity of applied measures (i.e. expressed as reduction of social contacts) and delay in the peak of new infections. As a consequence of this delay, measure intensity scales super-linearly with the required duration of application to show added value; i.e. a lower number of fatalities. Given the large scale of temporal delay (up to multiple years for a 10% increment of additional contact reduction), secondary effects of long-term social isolation such as psychological distress, depression [34], and increased mortality [35] are likely to unravel the added value of intense social distancing measures. This holds true for effective reproduction numbers above one. Below this threshold, an intensification of measures reduces the required duration of measure application.

In the absence of a vaccination, mitigation strategies are crucial to keep the number of severe and critical cases below the capabilities of the health care system. If the use of mitigation interventions is well balanced against capability limits, the time required to establish herd immunity linearly scales with available capabilities of the health care system (defined by the number of ICU beds in the simulation). Other important factors are the reproduction number and the severity of the disease (expressed by the fraction of cases that need ICU admission). Depending on the calibration of those factors, it is estimated that herd immunity on a national level will be established in more than 2 years from now. This is in line with an agent-based simulation study by [36], who indicate a duration of 2 years and 4 months for the Netherlands. According to a deterministic simulation by [16] in the United States the epidemic could last into 2022 under current critical care capabilities. Given this timescale, the success of a strategy based on social distancing, delay and herd immunity is unrealistic under known preconditions.

According to [37], an extinction strategy implemented by intense countermeasures seems promising. This is supported by our low effective reproduction scenario (*R* < 1).

If an extinction is not feasible, interventions should be as moderate as possible. Negative societal and economic consequences associated with drastic social-distancing are likely to undermine a required long-term measure application. The identified super-linear scaling (*R* > 1) between intensity and required duration of measure application does amplify this problem. As a result, countries whose policy is exclusively aimed at minimizing the number of cases are in danger of experiencing an untimely termination of countermeasures and a large second wave of outbreaks (compare model output in Fig. 7). Data on government response stringency [38] and daily confirmed deaths [39] reveal an association between intense measures and large second waves in countries like Albania, Bosnia and Herzegovina, Bulgaria, Egypt, Israel, Palestine, Romania or Serbia (data acquired at the end of August 2020). The probability that more resilient countries will experience similar problems rises every day without vaccination. So far (August 2020), countries like New Zealand or Vietnam did well in containing the virus by an aggressive elimination approach combined with comprehensive international travel restrictions [40] and by implementing timely, intense action [41] To date, a differentiated assessment of such ambitious policies is primarily constrained by the limited availability of studies on secondary effects of social distancing and isolation in the case of a global pandemic. Similarly, the assessment of more targeted countermeasures such as the selective isolation of vulnerable individuals or approaches of contact tracing and isolation are limited by data scarcity and in part data inconsistency.

For instance, there is little reliable information about age-stratified asymptomatic ratios. Moreover, the impact of country-based measures has hardly been empirically assessed by methods of inferential statistics. While such studies will shed light on important system dependencies, large-scale investment into health care and medical research is essential to spawn game-changing innovation such as the development of vaccines, drugs and affordable test kits.

## Data Availability

The data/parameters used in the manuscript are publicly available.

## References

1. Medicine JHU. Coronavirus COVID-19 Global Cases by the Center for Systems Science and Engineering (CSSE) at JHU [WWW Document]. online. 2020;.

2. Ng Y, Li Z, Chua YX, Chaw WL, Zhao Z, Er B, et al. Evaluation of the effectiveness of surveillance and containment measures for the first 100 patients with COVID-19 in Singapore-January. online. 2020;.

3. Parodi SM, Liu VX. From Containment to Mitigation of COVID-19 in the US. Jama. 2020;.

4. Ebrahim SH, Ahmed QA, Gozzer E, Schlagenhauf P, Memish ZA. Covid-19 and community mitigation strategies in a pandemic. British Medical Journal Publishing Group. 2020;.

5. Anderson RM, Heesterbeek H, Klinkenberg D, Hollingsworth TD. How will country-based mitigation measures influence the course of the COVID-19 epidemic? The Lancet. 2020;395:931.

6. Flaxman S, Mishra S, Gandy A, et al. Estimating the number of infections and the impact of non-pharmaceutical interventions on COVID-19 in 11 European countries. Imp Coll Prepr. 2020;.

7. Preiser W, van Zyl G, Dramowski A. COVID-19: Getting ahead of the epidemic curve by early implementation of social distancing. S Afr Med J. 2020;110:1.

8. Tuite A, Fisman DN, Greer AL. Mathematical modeling of COVID-19 transmission and mitigation strategies in the population of Ontario, Canada. medRxiv. 2020;.

9. Brooks SK, Webster RK, Smith LE, Woodland L, Wessely S, Greenberg N, et al. The psychological impact of quarantine and how to reduce it: rapid review of the evidence. The Lancet. 2020;.

10. Lugner AK, Postma MJ. Mitigation of pandemic influenza: review of cost-effectiveness studies. Expert Rev Pharmacoecon Outcomes Res. 2009;9:547.

11. Rhodes A, Ferdinande P, Flaatten H, Guidet B, Metnitz P, Moreno R. The variability of critical care bed numbers in Europe. Intensive Care Med. 2012;38:1647.

12. MacGregor H. Novelty and uncertainty: social science contributions to a response to COVID-19.;.

13. Kwakkel JH. The Exploratory Modeling Workbench: An open source toolkit for exploratory modeling, scenario discovery, and (multi-objective) robust decision making. Environ Model Softw. 2017;96:239.

14. Xu Z, Shi L, Wang Y, Zhang J, Huang L, Zhang C, et al. Pathological findings of COVID-19 associated with acute respiratory distress syndrome. Lancet Respir Med. 2020;.

15. Remuzzi A, Remuzzi G. COVID-19 and Italy: what next? The Lancet. 2020;.

16. Kissler SM, Tedijanto C, Lipsitch M, Grad Y. Social distancing strategies for curbing the COVID-19 epidemic. medRxiv. 2020;.

17. Shen M, Peng Z, Xiao Y, Zhang L. Modelling the epidemic trend of the 2019 novel coronavirus outbreak in China. bioRxiv. 2020;.

18. Huynh TLD. Does culture matter social distancing under the COVID-19 pandemic? Safety Science. 2020;104872.

19. Mizumoto K, Kagaya K, Zarebski A, Chowell G. Estimating the asymptomatic proportion of coronavirus disease 2019 (COVID-19) cases on board the Diamond Princess cruise ship, Yokohama, Japan. Eurosurveillance. 2020;25:2000180.

20. Sutton D, Fuchs K, Dalton M, Goffman D. Universal screening for SARS-CoV-2 in women admitted for delivery. New England Journal of Medicine. 2020;382.

21. Rocklov J. COVID-19 health care demand and mortality in Sweden in response to non-pharmaceutical (NPIs) mitigation and suppression scenarios. medRxiv. 2020;.

22. Bai Y, Yao L, Wei T, Tian F, Jin DY, Chen L, et al. Presumed asymptomatic carrier transmission of COVID-19. Jama. 2020;.

23. Guan W, Ni Z, Hu Y, Liang W, Ou C, He J, et al. Clinical characteristics of 2019 novel coronavirus infection in China. MedRxiv. 2020;.

24. Organization WWH. Health Systems Respond to COVID-19 - Creating surge capacity for acute and intensive care Recommendations for the WHO European Region. Technical Guidance WHO. 2020;2.

25. Grasselli G, Pesenti A, Cecconi M. Critical care utilization for the COVID-19 outbreak in Lombardy, Italy: early experience and forecast during an emergency response. Jama. 2020;.

26. Hofmarcher M, Singhuber C. Schutzschirm für das Gesundheitswesen in Zeiten von COVID-19. health systems intelligence. 2020;.

27. Onder G, Rezza G, Brusaferro S. Case-fatality rate and characteristics of patients dying in relation to COVID-19 in Italy. Jama. 2020;.

28. Shim E, Tariq A, Choi W, Lee Y, Chowell G. Transmission potential and severity of COVID-19 in South Korea. Int J Infect Dis. 2020;.

29. Khafaie MA, Rahim F. Cross-Country Comparison of Case Fatality Rates of COVID-19/SARS-COV-2. Osong Public Health Res Perspect. 2020;11:74.

30. Jung S, Akhmetzhanov AR, Hayashi K, Linton NM, Yang Y, Yuan B, et al. Real-time estimation of the risk of death from novel coronavirus (covid-19) infection: Inference using exported cases. J Clin Med. 2020;9:523.

31. Liu Y, Gayle AA, Wilder-Smith A, Rocklov J. The reproductive number of COVID-19 is higher compared to SARS coronavirus. J Travel Med. 2020;.

32. Koo JR, Cook AR, Park M, Sun Y, Sun H, Lim JT, et al. Interventions to mitigate early spread of SARS-CoV-2 in Singapore: a modelling study. Lancet Infect Dis. 2020;.

33. Dietz K. The estimation of the basic reproduction number for infectious diseases. Stat Methods Med Res. 1993;2:23.

34. Hawryluck L, Gold WL, Robinson S, Pogorski S, Galea S, Styra R. SARS control and psychological effects of quarantine, Toronto, Canada. Emerg Infect Dis. 2004;10:1206.

35. Cacioppo JT, Cacioppo S. Social relationships and health: The toxic effects of perceived social isolation. Soc Personal Psychol Compass. 2014;8:58.

36. de Vlas SJ, Coffeng LE. A phased lift of control: a practical strategy to achieve herd immunity against Covid-19 at the country level. medRxiv. 2020;.

37. Bock W, Adamik B, Bawiec M, Bezborodov V, Bodych M, Burgard JP, et al. Mitigation and herd immunity strategy for COVID-19 is likely to fail. medRxiv. 2020;.

38. Hale T, Webster S, Petherick A, Phillips T, Kira B. Oxford COVID-19 Government Response Tracker, Blavatnik School of Government. online. 2020;.

39. ECDC. Data on COVID-19 (coronavirus) by Our World in Data. online. 2020;.

40. Cousins S. New Zealand eliminates COVID-19. The Lancet. 2020;395.

41. Huynh TLD. The COVID-19 containment in Vietnam: What are we doing? Journal of Global Health. 2020;10.

